# Improved prostate cancer prediction by combining Prostate-Specific Antigen (PSA) test results with Genetic Risk Scores (GRS/PRS)

**DOI:** 10.64898/2026.05.14.26353195

**Authors:** Jingzhan Lu, Ge Chen, Samuel W. D. Merriel, Michael N. Weedon, Anna Murray, Sarah E. R. Bailey, Harry D. Green

## Abstract

**Background:** Prostate cancer is the second most common cancer in men worldwide. The Prostate Specific Antigen (PSA) blood test is widely used for prostate cancer detection but suffers from high false-positive rates (up to 80%). Genetic risk scores (GRS/PRS) have a similar performance to PSA testing in predicting prostate cancer risk.

**Method:** GRS_269_ for prostate cancer was derived using 269 known risk variants and applied to UK Biobank participants. We assessed whether GRS_269_ improved power to predict prostate cancer diagnosis on top of age and pre-prostatectomy PSA level among 17,380 cases. Longitudinal PSA measurements were processed as median, first, last (most recent), and random PSA. All models were adjusted for age.

**Results:** Across all PSA measures, the integrated model combining GRS_269_, PSA, and age consistently outperformed models using GRS_269_ or PSA alone. The highest predictive performance was observed using the last PSA value combined with GRS_269_ (AUC = 0.82, 95% CI: 0.81 - 0.82), compared to GRS_269_ alone (AUC = 0.70, 95% CI: 0.68 - 0.72) or PSA alone (AUC = 0.73, 95% CI: 0.70 - 0.75).

**Conclusion:** Combining genetic risk with PSA and age improves prostate cancer risk prediction in a population setting. These findings highlight the potential clinical implications of integrating GRS will enhance early prostate cancer prediction pathways in primary care.

## Introduction

Prostate cancer is the most common cancer in men in 112 countries [1] and the second leading cause of cancer death among men in the UK. One in eight UK men will be diagnosed with prostate cancer in their lifetime, and the disease is responsible for over 12,000 deaths annually [2]. Prostate cancer occurs predominantly in older men, with a median age of diagnosis being 67 years [3]. Prostate cancer typically progresses slowly, and many men die of competing causes [4]. This contributes to overdiagnosis and overtreatment in clinical settings [5]. Despite widespread use, the prostate-specific antigen (PSA) blood test has limited diagnostic accuracy, with a false-positive rate as high as 80% [6]; with an overall area under the receiver operator characteristic curve (AUC) for any prostate cancer of 0.72 (95% CI: 0.68 - 0.76) [7].

Prostate cancer has a strong genetic component, with heritability estimated at 58% in twin studies (95% CI: 52–63%) [8], emphasising the potential role of genetic risk assessment in improving risk stratification. Genetic risk scores (GRS), also referred to as polygenic risk scores (PRS), quantify an individual’s inherited susceptibility based on common genetic variants and are increasingly incorporated into cancer risk prediction models [9]. Incorporating genomics into routine clinical care is part of the UK NHS 10-year Plan [10]. As the NHS and global health systems move toward integrating population-based GRS into routine care, understanding how genetic information can augment existing diagnostic tools is critical. The incorporation of GRS into primary care pathways for suspected cancer has the potential to enhance risk stratification and support clinical decision-making at the point of consultation [11], [12].

Recent evidence from the BARCODE1 study, McHugh et al. (2025) further underscores the value of genetic risk stratification in prostate cancer screening. In the BARCODE1 study, screening men in the highest decile of genetic risk led to the detection of clinically significant prostate cancer, with 103 men diagnosed with intermediate- or high-risk disease. Notably, 74 of these cases (71.8%) would have been missed under current UK screening pathways based on PSA thresholds and MRI findings [13] BARCODE1 study demonstrated that GRS was more effective than PSA or MRI alone in identifying aggressive prostate cancer. In a separate study, GRS has been demonstrated to predict 2-year prostate cancer incidence with a similar performance to the PSA test in a symptomatic cohort [14]. The ongoing 15-year TRANSFORM trial aims to provide evidence on the clinical utility of GRSs in a more diverse population, but also plans to focus on men with a high GRS [15].

Building on these studies, our study leverages the full distribution of GRS within the UK Biobank (UKBB) cohort and includes PSA tests, enabling a broader assessment of how genetic risk and PSA levels interact across a large population. This approach allows for a more comprehensive evaluation of the utility of GRS in a potential screening project, particularly in improving early non-invasive risk stratification in a population setting. In this study, we investigated whether combining PSA test results with a GRS improves the prediction of prostate cancer compared to either measure alone.

KLK3 (kallikrein-related peptidase 3) is the functional protein known as PSA and serves as a key biomarker for the disease. Unlike clinical PSA records, these levels were obtained via the Olink high-throughput platform from a randomly selected 10% of UKBB participants [21]. By utilising this baseline subset, we aimed to validate whether the predictive synergy between GRS and PSA remains robust across the general population.

## Methodology

### Participants: UK Biobank (UKBB)

The UKBB is a prospective cohort study of people aged between 40 and 69 years across the United Kingdom that has collected in-depth phenotypic and genetic data on 488,377 participants and is linked with the electronic health records. Further information on UKBB, including genetic quality control procedures as described previously by Bycroft et al. (2018) [16]. All male participants in the UKBB with at least one available PSA test in linked primary care records were eligible for inclusion.

### GRS_269_Calculation

A genetic risk score (GRS_269_) for prostate cancer was constructed using 269 established risk variants identified in the largest trans-ancestry genome-wide association meta-analysis to date by Conti et al. (2021) [17], which included data from 107,247 prostate cancer cases and 127,006 controls across 136 studies contributed [18]. The UKBB data was used for validation but not for discovery. We used the log-odds ratio from the multi-ancestry SNP weighting to broaden the study population. These external weights were used instead of UKBB-specific estimates to minimise overfitting. The GRS_269_ for each UKBB participant was computed as the weighted sum of the genotype dosage for the 269 SNPs, representing the individual’s inherited genetic risk for prostate cancer.

### Variable definition

#### PSA testing

PSA levels typically fall to extremely low values within one month after prostate surgery; all PSA measurements recorded after surgery (where the PSA test age exceeded the surgery age) were excluded. To further ensure data integrity and minimise the influence of potential unrecorded surgical interventions, PSA values below 0.2 ng/mL were also removed, as this indicates recent surgery prior to biochemical recurrence [19], [20], and their presence in the UKBB likely reflects procedures missing from the OPCS-4 records. Additionally, extreme outliers above 500 ng/mL were excluded to improve data quality.

#### Median, First, Last and Random PSA testing

For each participant, PSA follow-up duration was defined as the time between the first eligible PSA measurement and the last measurement before prostate cancer diagnosis or prostate surgery. To compare alternative representations of longitudinal PSA data, four summary measures were derived from the preoperative records: the median PSA, the first (earliest round) PSA, the last (most recent round) PSA, and a randomly selected PSA value. For the random PSA measure, one value per participant was selected using a fixed random seed to ensure reproducibility.

#### Prostate Cancer and Controls Definition

In **Table 1**, Prostate cancer cases were identified using the ICD-10 code “*C61*” from the linked national cancer registry data. PSA test results were extracted from linked primary care records using the READ code “*43Z2*.*”* Cancer registry linkage is available in UKBB data until Oct. 2023, while GP record linkage is available until January 2022. Surgical interventions relevant to prostate disease were identified using OPCS-4 codes, including “*M34*.*1”* (cystoprostatectomy) and “*M65*.*1–M65*.*6”* (endoscopic resection of prostate). Genetically inferred male participants without a prostate cancer diagnosis were used as controls.

**Table 1.** Clinical Terminology and Case Definitions for Prostate Cancer in the UK Biobank Cohort.

| Description | Clinical Terminology | Codes |
| --- | --- | --- |
| **Primary Care** |  |  |
| Serum prostate specific antigen (PSA) level | Read Codes | 43Z2. |
| **Secondary Care** |  |  |
| Malignant neoplasm of prostate | ICD-10 code | C61 |
| Prostatectomy (Surgery) | OPCS-4 | M34.1, M65.1-M65.6 |

### Statistical Methods

All data processing and statistical analyses were performed using R version 4.4.0. Logistic regression models were used to evaluate the performance of PSA, GRS_269_, and their combination for prostate cancer. All models adjusted the age of the PSA test. Model discrimination was assessed using the area under the receiver (AUC) operating characteristic curve, which is a composite metric of sensitivity and specificity.

A heatmap was constructed to visualise the joint effects of PSA and GRS on prostate cancer risk. The x-axis represents GRS_269_ categories from the lowest 10% to the highest 100%, and the y-axis represents median PSA levels. Each cell shows the predicted probability of prostate cancer from the logistic regression model, with colour intensity ranging from white (lower risk) to dark blue (higher risk).

### Definition of outcomes for the Short-term and Overall Cancer Risk model

For a prostate cancer diagnosis within two years of the PSA test, it is likely that prostate cancer was already present at the point of testing, and the model is detecting pre-existing prostate cancer rather than predicting future disease. To evaluate model performance across different clinical time horizons, two prostate cancer outcome definitions were applied.

For short-term (≤2-year) risk, testing ability to detect existing cancer, cases were defined as individuals diagnosed with prostate cancer within two years of PSA measurement, with PSA values restricted to those obtained within this pre-diagnostic window. Controls were defined as individuals without a prostate cancer diagnosis within the same timeframe.

For overall risk, testing ability to predict future cancer onset, cases included all individuals diagnosed with prostate cancer during the full follow-up period, regardless of timing relative to PSA measurement, while controls comprised those without a recorded diagnosis throughout follow-up. Model performance was assessed separately under these two outcome definitions to compare discriminative ability across short-term detection and long-term risk prediction.

### KLK3 Sensitivity analysis

To address potential selection bias in GP-derived PSA records, where individuals undergoing testing may already be at higher clinical suspicion for prostate cancer, we conducted a sensitivity analysis utilising baseline KLK3 protein level. This analysis was restricted to a subset of 1,296 individuals who had both available GP-recorded PSA data and were included in the UKBB’s proteomics profiling. We assessed the discriminative performance of prediction models incorporating GRS_269_, PSA, age, and KLK3. Model performance was quantified using the AUC.

## Results

We identified 16,116 UKBB participants with a read code for Serum Prostate Specific Antigen (PSA) level, 17,380 with a cancer registry record for prostate cancer, and 9,446 with a record of prostatectomy surgery. Note that the PSA level results are restricted to individuals with linked primary care data (N~230,000), whereas prostate cancer and prostatectomy data are available in the full N~500,000 UKBB cohort.

Among 222,913 men in the UKBB of **Figure 1**, 16,116 had PSA measurements available from linked primary care records. After excluding post-prostatectomy PSA measurements, 13,504 individuals with pre-prostatectomy PSA data were retained. Of these, 13,218 men had complete genetic data and were eligible for analysis. Following exclusion of extreme PSA values (<0.2 ng/mL or >500 ng/mL), 4.8% of PSA records were excluded. The final analytical cohort comprised 12,574 men, including 1,187 prostate cancer cases identified through cancer registry records and 11,387 cancer-free controls.

**Figure 1.**
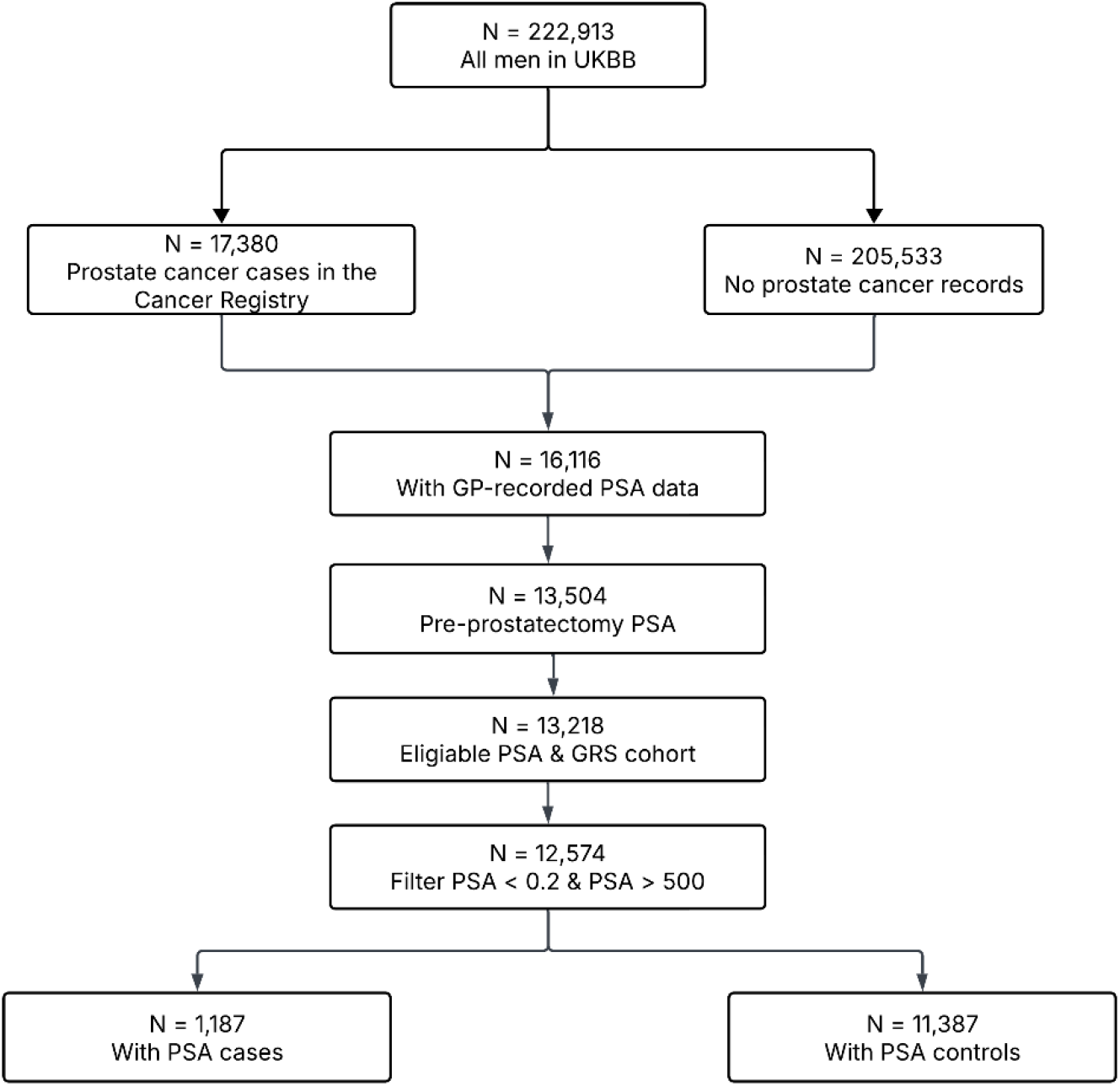
Flow diagram of the derivation of the PSA and GRS study cohort in the UK Biobank

The cohort characteristics stratified by case/control status are outlined in **Table 2**. Cases were significantly older (P < 2 × 10^−16^), had a higher PSA (P < 2 × 10^−16^) and GRS_269_ (P < 2 × 10^−16^), but deprivation and ethnicity were not significantly associated with case control status after adjusting for multiple testing.

**Table 2.** Observational associations between prostate cancer cases and controls. All odds ratios and p-values were estimated with logistic regression, with the exception of ethnicity, for which a Chi-squared test was performed.

|  | Cases | Controls | Odds ratio | P value |
| --- | --- | --- | --- | --- |
| **Number of participants** | 1,187 | 11,387 |  |  |
| **Mean age** | 66.27 ± 6.03 | 62.01 ± 7.48 | 1.09 | <2e-16 |
| **Median PSA** | 6.73 ± 19.06 | 1.88 ± 2.63 | 1.26 | <2e-16 |
| **Median GRS<sub>269</sub>** | -0.96 ± 0.71 | -1.48 ± 0.73 | 2.65 | <2e-16 |
| **Deprivation** | -1.90 ± 2.80 | -1.70 ± 2.93 | 0.98 | 0.02 |
| **Ethnicity** | 97% White | 97% White | P = 0.002 |  |
|  | 1% Asian | 1% Asian |  |  |
|  | 1% Black | 1% Black |  |  |

### PSA kinetics

In men with PSA data, 70.4% of participants had a follow-up duration of less than five years, defined as the time span between their first and last PSA tests. This relatively short follow-up period helps to minimise age-related biological variation in PSA levels. Additionally, 83.4% of participants had fewer than five PSA measurements recorded; therefore, the individual median PSA value was calculated to provide a stable summary measure for each participant. To further address potential misclassification due to unrecorded surgery, which likely reflects this threshold-based filtering improved the best predictive performance with AUC 0.78 (95% CI: 0.76-0.79), (compare with AUC = 0.73, 95% CI: 0.71-0.75, using raw PSA levels), as shown in **Figure 2**.

**Figure 2.**
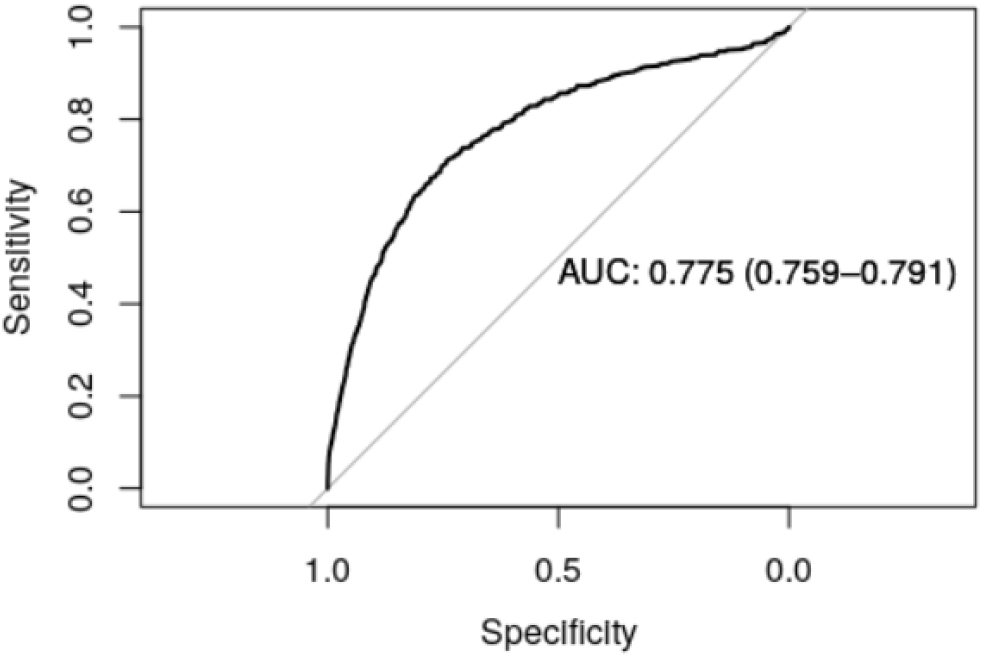
ROC curve showing improved prostate cancer risk prediction when excluding PSA values <0.2 ng/mL) model.

### GRS Category and Median PSA value

This heatmap of **Figure 3** illustrates how PSA and genetic risk jointly influence the likelihood of prostate cancer. Individuals with high PSA but low GRS, as well as those with high GRS despite having low PSA, both show elevated predicted risk.

**Figure 3.**
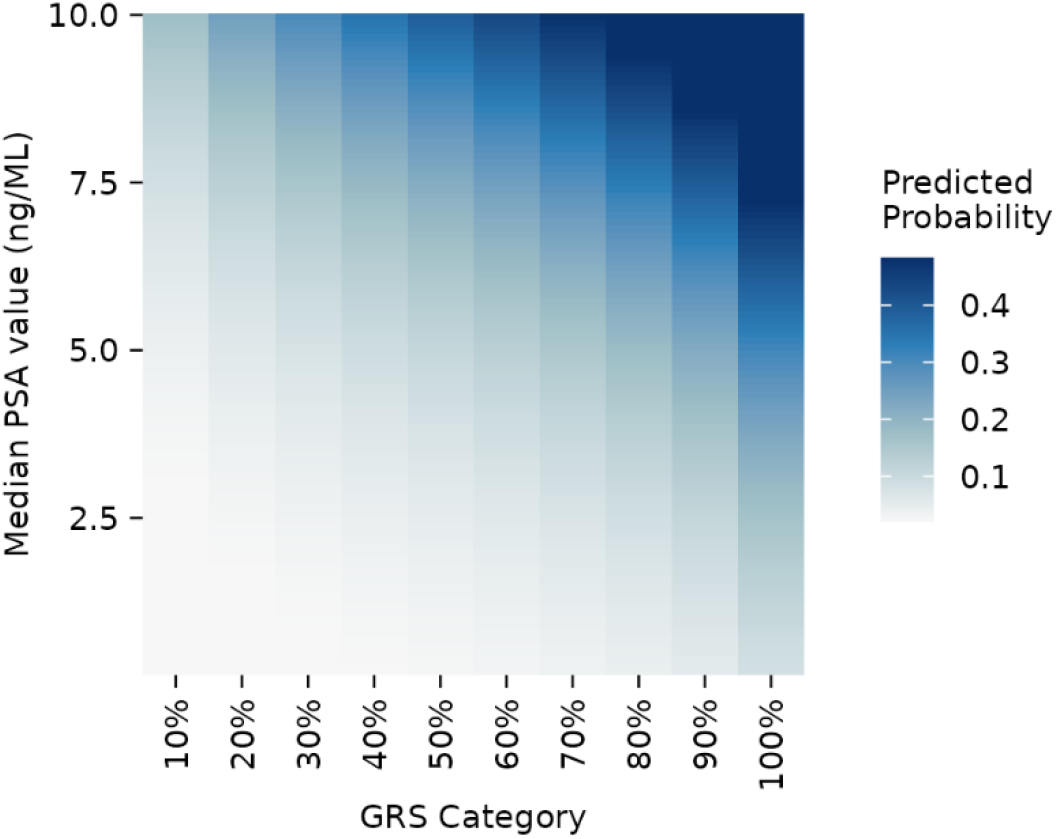
Predicted prostate cancer probability across GRS categories and median PSA levels

Across all four selection PSA criteria: (a) Median, (b) First, (c) Last, and (d) Random, the full model incorporating GRS_269_, PSA, and age consistently outperformed models using GRS_269_ or PSA alone (Table 2). For (a) Median, the full model achieved the highest discrimination with an AUC of 0.81, compared with 0.75 for GRS_269_ + age and 0.77 for PSA + age. Using (b) First, the full model reached an AUC of 0.78, while GRS_269_ + age and PSA + age yielded 0.74 and 0.71. Performance peaked at an AUC of 0.82 with (c) Last, compared with 0.76 and 0.77 for the GRS_269_ + age and PSA + age models. Similarly, with (d) Random, the full model maintained strong performance (AUC = 0.81), highlighting the robustness of the combined approach across different PSA measures.

**Table 3.** Comparison of AUC values across PSA measurement strategies (Median, First, Random, Last) showing consistent improvement with the full model (GRS_269_ + PSA + Age).

| Models * | Median PSA | First PSA | Last/Recent PSA | Random PSA |
| --- | --- | --- | --- | --- |
| GRS<sub>269</sub> | 0.70 (0.68-0.72) | - | - | - |
| Age | 0.66 (0.65-0.68) | 0.61 (0.60-0.62) | 0.68 (0.67-0.70) | 0.65 (0.64-0.67) |
| PSA | **0.75 (0.73-0.77)** | **0.72 (0.70-0.74)** | **0.73 (0.70-0.75)** | **0.77 (0.75-0.79)** |
| GRS<sub>269</sub> + Age | 0.75 (0.74-0.77) | 0.74 (0.72-0.75) | 0.76 (0.75-0.78) | 0.75 (0.73-0.76) |
| GRS<sub>269</sub> + PSA | **0.79 (0.79-0.81)** | **0.77 (0.76-0.78)** | **0.80 (0.78-0.81)** | **0.80 (0.78-0.81)** |
| PSA + Age | 0.77 (0.76-0.79) | 0.71 (0.70-0.73) | 0.77 (0.75-0.78) | 0.76 (0.74-0.78) |
| GRS<sub>269</sub> + PSA + Age | **0.81 (0.80-0.83)** | **0.78 (0.77-0.79)** | **0.82 (0.81-0.82)** | **0.81 (0.80-0.83)** |

\*Logistic Regression model results: AUC (95% CI)

Using the most recent (last) PSA measurement, the full model incorporating GRS_269_, PSA, and age achieved the highest predictive performance, with an AUC of 0.82 (95% CI: 0.81–0.82). These results suggest that this approach may be particularly informative for quantifying risk in later life.

### Model Performance for Short-term and Overall cancer risk

The accuracy of short-term (≤2-year) detection and overall prostate cancer prediction across the full follow-up period is summarised in Supplementary Table 1. Overall, the models demonstrate consistently strong performance for short-term risk, with AUCs generally higher than or comparable to those observed for overall risk. In particular, the integrated model combining GRS_269,_ PSA and Age achieve the highest discriminative ability for both outcomes (AUC 0.84, 95% CI: 0.83–0.86 for ≤2-year detection; AUC 0.81, 95% CI: 0.80–0.83 for overall prediction). This suggests that integrating genetic risk (GRS_269_) with recent PSA levels provides robust short-term risk stratification, while maintaining strong predictive performance across the entire follow-up period.

### KLK3 Sensitivity analysis

In the sensitivity analysis, models incorporating KLK3 demonstrated consistent and robust discriminative performance (Supplementary Table 2). The combined model including GRS269, PSA, age, and KLK3 achieved the highest AUC of 0.80 (95% CI: 0.76– 0.84), while models including KLK3 alone or in combination with age also showed strong predictive ability (AUC up to 0.75, 95% CI: 0.70–0.80). Overall, these results were comparable to the primary analyses, indicating stable model performance.

## Discussion

### Key Findings

Our findings show that combining genetic risk scores (GRS_269_) with prostate-specific antigen (PSA) testing improves prostate cancer prediction compared with using either marker alone. PSA levels fluctuate over time and require repeated measurements, whereas GRS remains largely stable across the lifespan and only needs to be assessed once, providing a simple and durable tool for risk stratification in clinical practice. These results are consistent with a study from Schaffer et al. (2023), which included 655 African- and European-ancestry men undergoing prostate biopsy [21]. In that cohort, the GRS_269_ adjusted for age achieved an AUC of 0.70 for predicting any prostate cancer, compared with 0.66 for PSA + age, while combining PSA, GRS_269_ and age improved the AUC to 0.72. Although the improvement for higher-grade aggressive prostate cancer (from biopsy) was minimal, the overall findings reinforce that integrating genetic risk information with PSA enhances predictive performance for prostate cancer.

Notably, the models demonstrated strong discriminative performance for short-term (≤2-year) risk, with the integrated model achieving particularly high accuracy. This indicates that integrating genetic risk (GRS_269_) with recent PSA levels and age provides effective short-term risk stratification, supporting the clinical utility of these models for the detection of existing prostate cancer.

### Strengths and Limitations

Our study uses the UKBB cohort, which provides a large, well-characterised population with longitudinal PSA measurements and genomic data. This enabled us to systematically evaluate multiple representations of PSA, including median, first, last (most recent) and randomly selected values, and to assess their predictive performance in combination with GRS. UKBB is also one of the only cohorts linking full genome sequencing with cancer registry and routinely collected primary care data, allowing estimation of the potential benefit for integrating cancer GRS into risk stratification in primary care. To our knowledge, this is the first study to directly compare these PSA measurement strategies and GRS at the same time, providing new insights into how PSA dynamics and genetic risk can be integrated to improve prostate cancer prediction.

PSA testing data in this study were derived from primary care records, which may introduce potential selection bias, as individuals undergoing PSA testing in a primary care setting are often at higher risk for prostate cancer. However, our sensitivity analyses using KLK3 suggest that the primary findings are robust against such bias, reinforcing the reliability of our integrated risk prediction approach. While the KLK3-based analysis (as demonstrated by [7]) shows strong potential for prostate cancer risk stratification, its clinical adoption is currently limited by high analytical costs and the need for specialised laboratory infrastructure. Therefore, we maintain that PSA remains the more cost-effective and scalable biomarker for population-based risk stratification in routine clinical practice.

This study is also limited by the lack of detailed information on the stage and grade of prostate cancer in the UKBB, restricting our ability to fully evaluate GRS performance for high-risk aggressive cases. Additionally, the UKBB cohort is not fully representative of the general population and is enriched for healthy volunteers. This is especially true of participants with a PSA test in GP records, as 97% of our cohort is from a white ethnic background. The 269 variant prostate cancer GRS used in this study performs best in individuals of the European population and is known to underperform in individuals of the African population, who also have the highest background prostate cancer risk, so care must be taken to not widen existing healthcare disparities. More recently developed prostate cancer GRS perform better in multi-ancestry populations and should continue to be evaluated for more equitable application of GRS for cancer risk stratification [22].

### Implications

As genotyping costs continue to fall, GRS become an increasingly feasible addition to clinical practice [23], [24]. Risk stratification using both PSA and GRS could more accurately identify patients who could benefit from further testing, including prostate MRI and biopsy, even when only one of the two indicators is abnormal. This could facilitate more effective use of confirmatory diagnostic tests, which can be painful and carry risks of infection and complications [25].

With large-scale biobank resources such as Our Future Health [26] and All of US [27], the development of integrated risk tools is becoming increasingly feasible. The availability of genotype data in these datasets also makes it easier to calculate individual GRS directly, facilitating personalised risk prediction for prostate cancer. CanRisk-Prostate is an adaptation designed for prostate cancer that integrates clinical and demographic data [12] and is being evaluated for wider adoption into clinical practice.

The UK National Screening Committee (UK NSC) are currently reevaluating their approach to prostate cancer screening. Our results demonstrate an alternative approach to identifying high-risk men who may be at increased risk of prostate cancer, and who may benefit from earlier intervention.

## Acknowledgements

We would like to thank the Higgins family, Devonshire Provincial Grand Chapter and the Royal Arch Freemasons Devonshire for their generous donations, which supported this work. SERB is supported by a National Institute for Health and Care Research (NIHR) Advanced Fellowship (NIHR301666). SWDM is supported by the National Institute for Health and Care Research (NIHR) Manchester Biomedical Research Centre (BRC) (NIHR203308). This study was also funded by the National Institute for Health and Care Research Exeter Biomedical Research Centre (NIHR Exeter BRC) and the University of Exeter Medical School.

## Additional Information

### Authors’ contributions

HG and SERB formulated the initial hypothesis. JL, GC, SWDM, MW, AM, SERB, and HG designed the study. JZ carried out data preparation and analysis. All authors reviewed the results and participated in their interpretation. JZ and HG drafted the manuscript. GC and SWDM organised the dataset. All authors contributed to the final manuscript. SERB and HG are the guarantors. All authors take the responsibility for the decision to submit the manuscript. The corresponding author declares that all listed authors meet the authorship criteria and that no eligible author has been omitted.

### Ethics approval and consent to participate

This research has been conducted using the UK Biobank Resource under application number 103356. The ethics approval for UKBB was obtained from the North West Centre for Research Ethics Committee (11/NW/0382); all participants provided informed consent for research use.

### Consent for publication

Not applicable.

### Data availability

All data in this project were obtained from the UK Biobank resource, application number 103356. For information on accessing the UK Biobank, please visit https://www.ukbiobank.ac.uk/enable-your-research/apply-for-access.

### Code availability

All code used to generate the findings of this study is available on the author’s GitHub page: https://github.com/Jingzhan-Lu/GRS_PSA_UKBB. The GRS calculations and UKBB Health Records employed the following R functions, developed by HG and BMR at the

University of Exeter, for analysing UKBB genetic data within the DNA Nexus RStudio Workbench implementation: https://github.com/hdg204/UKBB.

