## Supplementary Material for "Improved prostate cancer prediction by combining Prostate-Specific Antigen (PSA) test results with Genetic Risk Scores (GRS/PRS)"

**Supplementary Table 1.** Discriminative performance (AUC) of prediction models for short-term (≤2-year) detection versus overall prediction prostate cancer risk.

| Models | Short-term Detection  AUC (95% CI) | Overall Prediction  AUC (95% CI) |
| --- | --- | --- |
| GRS_269_ | 0.71 (0.69-0.73) | 0.70 (0.68-0.72) |
| Age | 0.71 (0.70-0.73) | 0.66 (0.65-0.68) |
| PSA | 0.73 (0.71-0.75) | 0.75 (0.73-0.77) |
| GRS_269_ + Age | 0.79 (0.78-0.81) | 0.75 (0.74-0.77) |
| GRS_269_ + PSA | 0.81 (0.79-0.82) | 0.79 (0.79-0.81) |
| PSA + Age | 0.79 (0.78-0.81) | 0.77 (0.76-0.79) |
| GRS_269_ + PSA + Age | 0.84 (0.83-0.86) | 0.81 (0.80-0.83) |

**Supplementary Table 2.** Sensitivity analysis using KLK3 levels (N=1,296) to validate the predictive performance of risk models.

| Models | Short-term detection  AUC (95% CI) | Overall Prediction  AUC (95% CI) |
| --- | --- | --- |
| GRS_269_ | 0.71(0.64-0.78) | 0.69 (0.64 - 0.74) |
| PSA | 0.73(0.65-0.81) | 0.65 (0.59 - 0.72) |
| KLK3 | 0.78(0.72-0.84) | 0.74 (0.69 - 0.79) |
| GRS_269_ + KLK3 | 0.82(0.78-0.87) | 0.78 (0.74 – 0.82) |
| GRS_269_ + PSA | 0.78(0.72-0.83) | 0.73 (0.69 - 0.78) |
| GRS_269_ + PSA + KLK3 | 0.84(0.79-0.88) | 0.75 (0.70 - 0.80) |
| PSA + Age | 0.75(0.70-0.81) | 0.69 (0.64 - 0.73) |
| KLK3 + Age | 0.81 (0.75-0.86) | 0.75 (0.70 - 0.80) |
| GRS_269_ + KLK3 + Age | 0.86 (0.82-0.90) | 0.79 (0.75 – 0.83) |
| GRS_269_ + PSA + Age | 0.83 (0.79-0.88) | 0.77 (0.73 - 0.81) |
| GRS_269_ + PSA + Age + KLK3 | 0.87 (0.83-0.91) | 0.80 (0.76 - 0.84) |

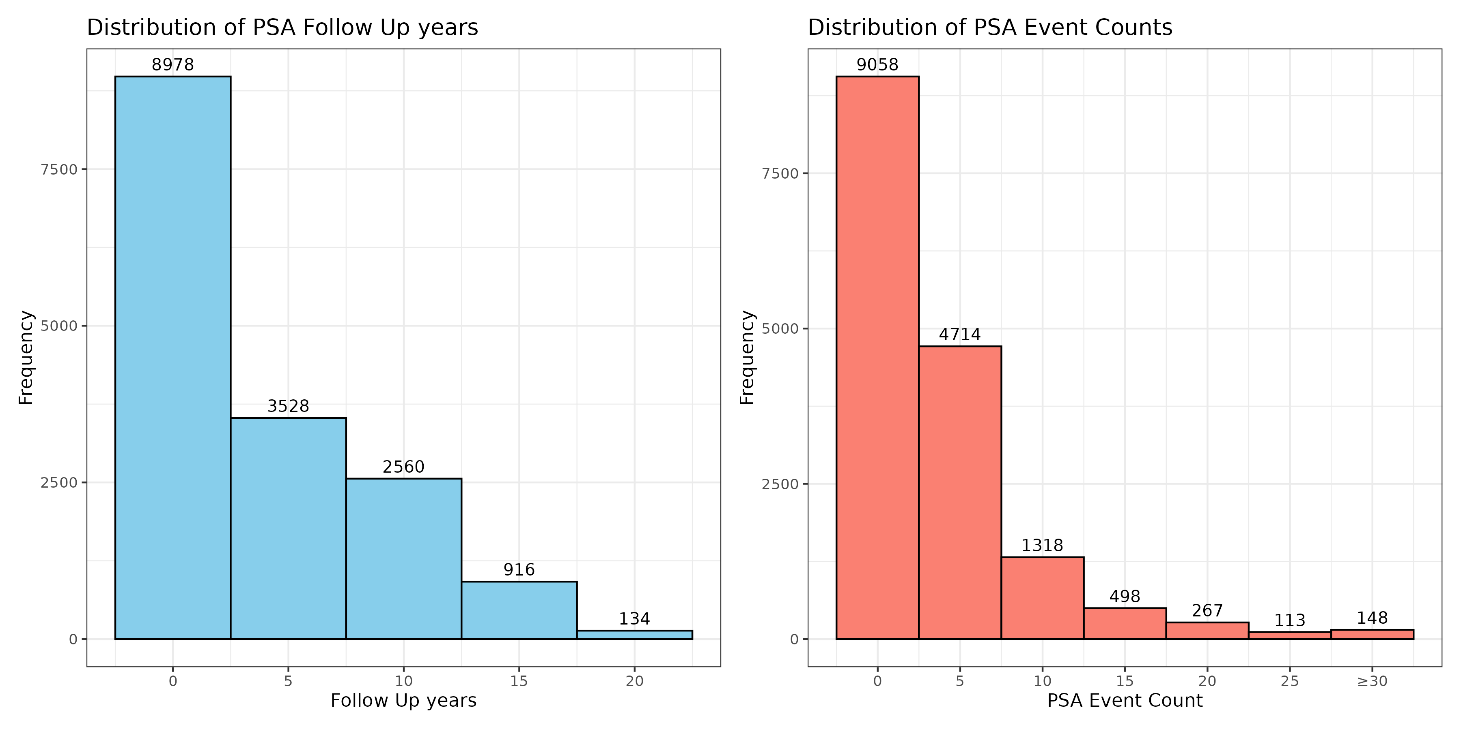

**Supplementary Figure 1.** PSA testing characteristics among UK Biobank participants. (A) Distribution of PSA follow-up duration (years) per individual, showing 70.4% had follow-up less than five years. (B) Distribution of the number of PSA test records per individual, with 83.4% having fewer than five tests.

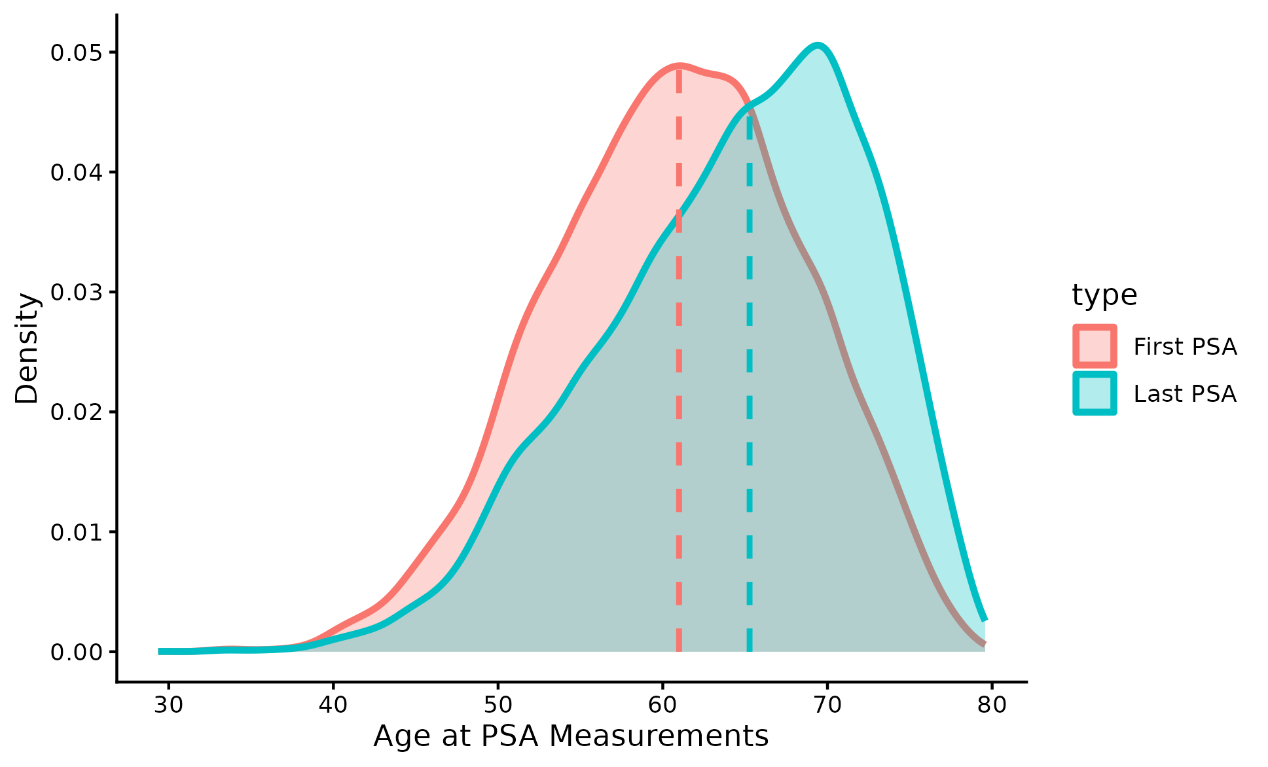

**Supplementary Figure 2.** Distribution of patient age at events in the first and final PSA models.

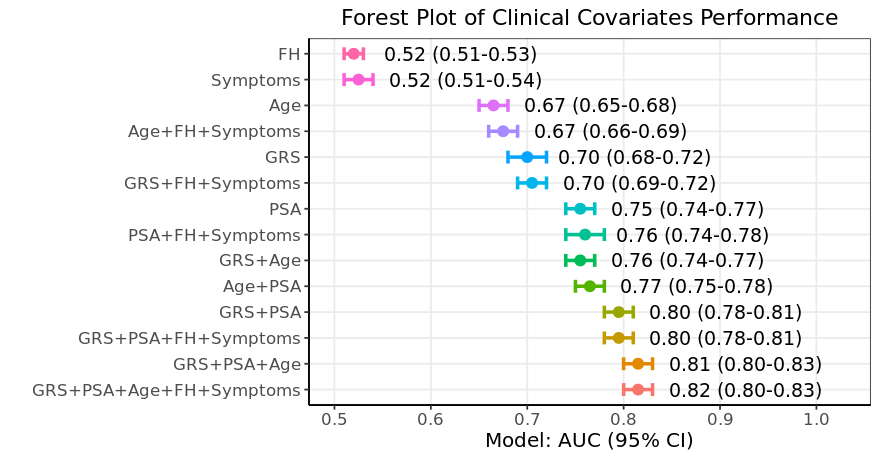

**Supplementary Figure 3.** Forest plot comparing AUC values across models with and without inclusion of family history and symptom data.

Family history and symptoms do not improve predictive accuracy over GRS and age: Incorporating prostate cancer or breast cancer family history and LUTS symptom information did not substantially improve predictive performance beyond GRS and age. As shown in the forest plot, the full model (GRS + PSA + age) achieved an AUC of 0.81, which increased only marginally to 0.82 when family history and symptoms were included.
